# Towards Medical Billing Automation: NLP for Outpatient Clinician Note Classification

**DOI:** 10.1101/2023.07.07.23292367

**Authors:** Matthew G. Crowson, Emily Alsentzer, Julie Fiskio, David W. Bates

**Affiliations:** Department of Otolaryngology-Head & Neck Surgery, Massachusetts Eye & Ear, Boston, Massachusetts, USA; Department of Otolaryngology-Head & Neck Surgery, Harvard Medical School, Massachusetts, USA; Division of General Internal Medicine and Primary Care, Brigham and Women’s Hospital, Boston, MA, USA; Department of Health Policy and Management, Harvard T. H. Chan School of Public Health, Boston, MA, USA

**Keywords:** natural language processing, provider billing, level of service, outpatient care

## Abstract

**Objectives:** Our primary objective was to develop a natural language processing approach that accurately predicts outpatient Evaluation and Management (E/M) level of service (LoS) codes using clinicians’ notes from a health system electronic health record. A secondary objective was to investigate the impact of clinic note de-identification on document classification performance.

**Methods:** We used retrospective outpatient office clinic notes from four medical and surgical specialties. Classification models were fine-tuned on the clinic notes datasets and stratified by subspecialty. The success criteria for the classification tasks were the classification accuracy and F1-scores on internal test data. For the secondary objective, the dataset was de-identified using Named Entity Recognition (NER) to remove protected health information (PHI), and models were retrained.

**Results:** The models demonstrated similar predictive performance across different specialties, except for internal medicine, which had the lowest classification accuracy across all model architectures. The models trained on the entire note corpus achieved an E/M LoS CPT code classification accuracy of 74.8% (CI 95: 74.1-75.6). However, the de-identified note corpus showed a markedly lower classification accuracy of 48.2% (CI 95: 47.7-48.6) compared to the model trained on the identified notes.

**Conclusion:** The study demonstrates the potential of NLP-based document classifiers to accurately predict E/M LoS CPT codes using clinical notes from various medical and procedural specialties. The models’ performance suggests that the classification task’s complexity merits further investigation. The de-identification experiment demonstrated that de-identification may negatively impact classifier performance. Further research is needed to validate the performance of our NLP classifiers in different healthcare settings and patient populations and to investigate the potential implications of de-identification on model performance.

## INTRODUCTION

The administrative burden of clinical billing activities for health insurance reimbursement is substantial and contributes to rising healthcare costs in the United States and other insurance-based systems worldwide ^1, 2^. Compared to other western countries, the United States’ proportion of total hospital costs devoted to administrative tasks is higher--it has exceeded 25% and has been rising ^2^. Administrative tasks related specifically to billing and insurance cost the United States healthcare system an estimated $471 billion in 2012, comprising nearly 15% of all healthcare spending ^3^. At the institutional level, the estimated billing and insurance-related administration costs range widely from $20 for a primary care visit to $215 for an inpatient surgical procedure ^4^.

Several reasons for the administrative cost burden have been proposed, but the complexity of the United States’ healthcare reimbursement scheme appears to be the leading cause ^2, 5^. At the individual practice level, physicians and clinic team members devote considerable time interacting with health plans daily ^6, 7^. Proposed cost mitigation efforts have ranged from reforming healthcare payment processes via standardizing payment rules, claim forms, and other innovations to computer-assisted claims processing systems that automate clinicians’ receipt, validation, formatting, and sending of individual claims in an attempt to reduce the time for completion ^8, 9^.

Natural language processing (NLP) is a subfield of artificial intelligence devoted to understanding and analyzing human language. It has seen exponential interest in applications across all domains and subspecialties of medicine ^10, 11^. NLP has been used for a variety of tasks in medicine spanning pharmaceutical and biological knowledge discovery, adverse event detection, prognosis modeling, clinical document classification, decision support systems, and point-of-care assistive technologies such as patient-facing chatbots and triage tools ^10, 12–17^. The advantages of NLP approaches include unlocking unstructured data types in clinical narratives and incorporating alternative data sources in diagnostic or prognostic predictive models. Transformer-based NLP models have demonstrated large improvement gains over several tasks, and open-source NLP libraries have made their implementation practical. Recent work has demonstrated the utility of transformer NLP models for classifying medical entities ^18, 19^, including diagnosis codes (e.g., International Classification of Diseases (ICD) codes) ^20, 21^, as well as Current Procedural Terminology (CPT) codes from clinical pathology reports ^22^. However, to the best of our knowledge, no work has leveraged NLP models to classify Evaluation and Management (E/M) level of service (LoS) codes. Healthcare providers use outpatient E/M LoS codes to report the complexity and intensity of patient visits for billing and reimbursement purposes. An accurate point-of-care clinical note LoS decision support tool may help increase administrative efficiency and reduce variation in clinical encounter coding.

In this work, we developed an NLP-based classifier to predict the Evaluation and Management (E/M) level of service (LoS) codes from outpatient clinician notes. We compared the performance of several general-domain and clinical language models and assessed the impact of fine-tuning on specialty-specific notes. Furthermore, we de-identified the clinical notes and studied the impact of note de-identification on classifier importance. This study demonstrates that NLP-based document classifiers can accurately predict E/M LoS CPT codes using clinical notes from various medical and procedural specialties. Further development of this autonomous classification approach might contribute to redesigning administrative billing processes to reduce resource and cost burden.

## RESULTS

### Clinic Notes Included in Modeling

Individual notes lacking a ground-truth label were removed, and notes with a E/M LoS CPT ‘Level I’ code were removed before model training as these codes represented less than 1% of all records (**Table 2**). Most notes had either Level III or Level IV coding. After data preprocessing, 31,115 patient notes were available for model development across all subspecialties (Cardiology, n =13,279; Gastroenterology, n = 5,299; Internal Medicine, n =10,178; Otolaryngology-Head & Neck Surgery, n = 2,197). The E/M LoS CPT ‘new patient’ codes distributions differed across specialties (**Table 2**).

**Table 1.** Mock example of de-identification process using named entity recognition of protected health information and obfuscation.

|  |  |
| --- | --- |
| NAME: **Earl Fullness**<br>MRN: **138582469**<br>DOB: **01/23/1987**<br>Date of service: **03/02/2023**<br>Location: **Mass Eye & Ear Hospital**<br>PCP: Dr. **B. Sick**<br>ASSESSMENT:<br>**35-year-old** male with a history of repeated left ear infections.<br>He has previously sought treatment at several hospitals in the **Boston** area including **Mass General Hospital** and **Brigham & Women's**. He reports experiencing pain and discomfort in his left ear, accompanied by redness, drainage, and decreased hearing. The patient has no other significant past medical history and takes no regular medications. No allergies are reported.<br>Further evaluation, including a physical examination and potentially imaging studies, is recommended to determine the cause of the persistent infections and to develop an appropriate treatment plan.<br>Provider: Dr. **N Cerumen**<br>Department: Otolaryngology-Head & Neck Surgery<br>Phone Number: | NAME: <NAME><br>MRN: <ID><br>DOB: <DATE><br>Date of service: <DATE><br>Location: <LOCATION><br>PCP: Dr. <NAME>:<br>ASSESSMENT:<br><AGE> male with a history of repeated left ear infections.<br>He has previously sought treatment at several hospitals in the <LOCATION> area including <LOCATION> and <LOCATION>.<br>He reports experiencing pain and discomfort in his left ear, accompanied by redness, drainage, and decreased hearing. The patient has no other significant past medical history and takes no regular medications. No allergies are reported.<br>Further evaluation, including a physical examination and potentially imaging studies, is recommended to determine the cause of the persistent infections and to develop an appropriate treatment plan.<br>Provider: Dr. <NAME><br>Department: Otolaryngology-Head & Neck Surgery<br>Phone Number: <CONTACT> |

**Table 2.**
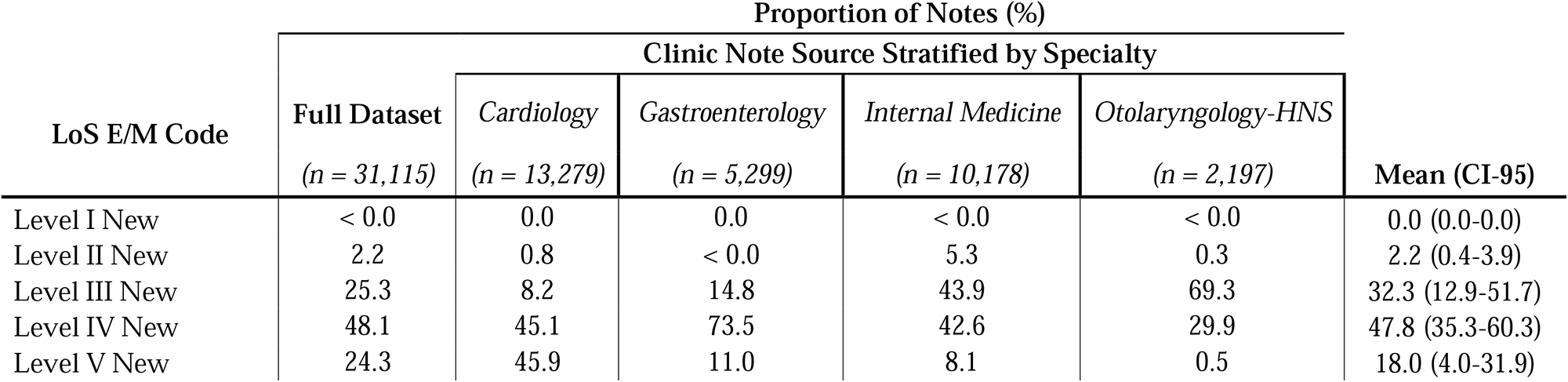
Distribution of Evaluation and Management (E/M) level of service (LoS) codes across different specialties included in modeling.

### Model Performance

The models trained on the entire note corpus achieved an E/M LoS CPT code classification accuracy of 75.6% (classification accuracy CI 95: 73.0-78.3%; weighted F1-score CI-95: 0.73-0.78). The predictive performance of the models trained on specialty-specific data was similar (**Table 3**; **Table 4**).

**Table 3.**
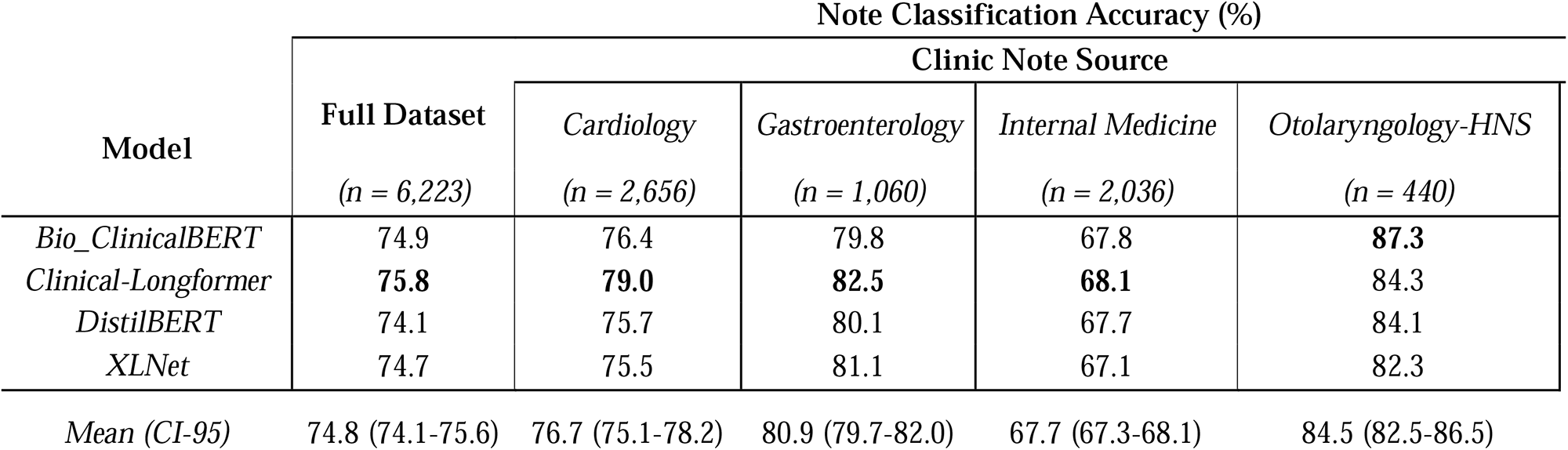
LoS E/M Code classification accuracy for general domain and clinical language models finetuned on notes from different medical specialties. All metrics are reported on test set data.

| Model | Note Classification Accuracy (%) |  |  |  |  |
| --- | --- | --- | --- | --- | --- |
|  | Full Dataset<br>(*n* = 6,223) | Clinic Note Source |  |  |  |
|  |  | Cardiology<br>(*n* = 2,656) | Gastroenterology<br>(*n* = 1,060) | Internal Medicine<br>(*n* = 2,036) | Otolaryngology-HNS<br>(*n* = 440) |
| *Bio_ClinicalBERT* | 74.9 | 76.4 | 79.8 | 67.8 | **87.3** |
| *Clinical-Longformer* | **75.8** | **79.0** | **82.5** | **68.1** | 84.3 |
| *DistilBERT* | 74.1 | 75.7 | 80.1 | 67.7 | 84.1 |
| *XLNet* | 74.7 | 75.5 | 81.1 | 67.1 | 82.3 |
| *Mean (CI-95)* | 74.8 (74.1-75.6) | 76.7 (75.1-78.2) | 80.9 (79.7-82.0) | 67.7 (67.3-68.1) | 84.5 (82.5-86.5) |

**Table 4.**
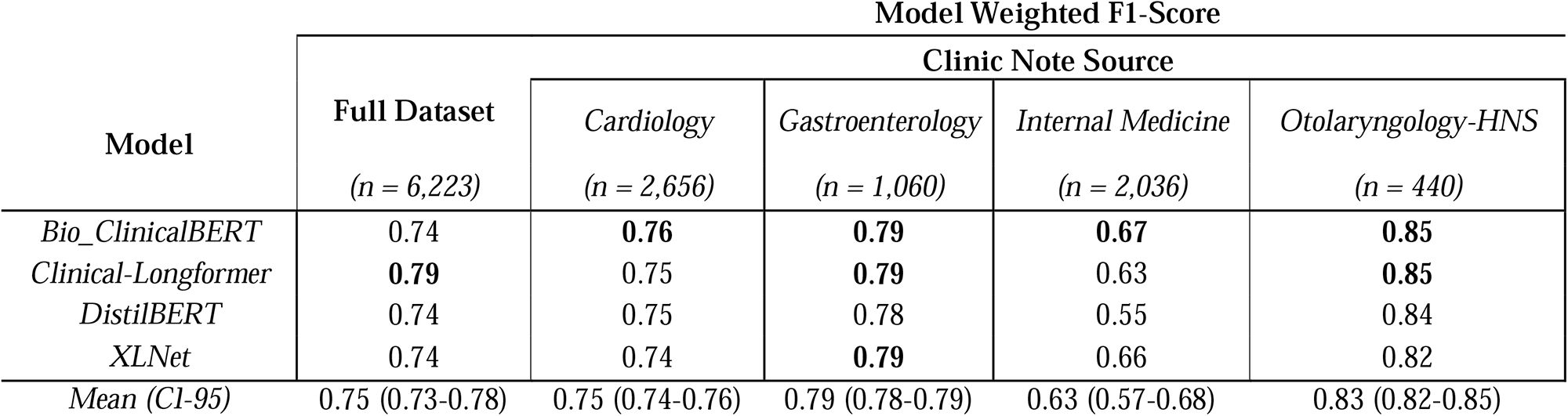
Weighted F1-scores for prediction of LoS E/M codes across general domain and clinical language models finetuned on notes from different medical specialties. All metrics are reported on test set data.

Internal medicine had the lowest classification accuracy (CI-95: 66.0 to 68.0%; weighted F1-score CI-95: 0.57-0.68), whereas Otolaryngology-Head & Neck surgery had the highest (classification accuracy CI-95: 82.3 to 84.6%; weighted F1-score CI-95: 0.82-0.85). Within each specialty, the models do not substantially differ in predictive performance (**Table 3**; **Table 4**). However, *Clinical-Longformer* generally had a higher predictive performance than the other models. The models demonstrated variation in performance across E/M LoS classes, likely due to class imbalance in the data (**Supplemental Tables S1-S5**). The models trained on the de-identified note corpus achieved an E/M LoS CPT code classification accuracy of 48.3% on average (classification accuracy CI 95: 48.0-48.5; **Table 5**).

**Table 5.** De-identified note classification performance for models trained on all clinic notes. All metrics are reported on test set data.

| **Model** | **LoS Classification**<br>**Accuracy (%)** | **Weighted F1-**<br>**Score** |
| --- | --- | --- |
|  | *Full Dataset (n = 6,223)* |  |
| *Bio_ClinicalBERT* | 48.4 | 0.45 |
| *Clinical-Longformer* | 47.9 | 0.42 |
| *DistilBERT* | 48.3 | 0.43 |
| *XLNet* | 48.1 | 0.44 |
| *Mean (CI-95)* | 48.3 (48.0-48.5) | 0.43 (0.42-0.45) |

## DISCUSSION

In this study, we developed natural language processing (NLP) document classifier models to assign Evaluation and Management (E/M) level of service (LoS) codes using clinical notes from an electronic health record. Our models that were trained on the entire clinical note corpus produced reasonable classification accuracy. The four models trained on the subspecialty notes showed similar predictive performance across different specialties, except for internal medicine, which had the lowest classification accuracy across all model architectures. Our results suggest that an NLP-based decision support tool for outpatient clinic notes might be a feasible approach with further development. Implementing such a tool might save healthcare system costs by reducing administrative burden and minimizing the potential for billing errors and fraud. Additionally, the tool could also alleviate the workload of physicians and clinic staff, allowing them to focus on patient care.

The overall classification accuracy of the model trained on the entire note corpus was good, indicating that our NLP-based approach might be useful in predicting E/M LoS CPT codes. However, we observed variability in classifier performance across models fine-tuned separately on notes from different medical specialties and across LoS CPT code levels. There could be several reasons for this observation. First, medical language is often characterized by complex terminology, abbreviations, and jargon that can be challenging for an NLP model to classify accurately. It is plausible that some medical specialties may have more complex or specialized language. Second, variability in documentation practices such as differences in documentation styles, structure, and terminology between specialties, could contribute to variation across specialties. This may be due, in part, to the diverse patient populations and conditions typically encountered within each specialty. Last, clinicians within medical specialties may have idiosyncratic standards for assigning E/M LoS codes. These possibilities have support in prior work that has demonstrated considerable variability in both the electronic health record utilization ^23^ and variation in electronic health record documentation between clinicians belonging to different medical specialties and health systems ^24^. Finally, there was class imbalance in the training data, which likely also contributed to performance variability. Taken together, the classifiers may struggle to generalize to these sources of variability, resulting in lower performance for some specialties versus others.

Across the clinical specialties, we observed that the ‘short-form’ models (i.e., models that accept 512 tokens) tended to underperform the *Clinical-Longformer* model in classification accuracy, which accepts inputs with a longer token length (i.e., a maximum token length of 4,096). The performance gap was not large. This finding is counterintuitive as we would expect longer notes to contain a richer representation and more informative features derived from the clinical encounter. One possible explanation is that the input sequences in clinical notes may contain redundant and irrelevant information, which might offset the benefits of having longer input sequences. Additionally, the pertinent and predictive information may be more likely to be contained in the first portion of the note’s body. This portion of a standard clinical note tends to be occupied by the patient’s chief complaint, history, and physical examination.

Clinicians contribute to the rising administrative cost of billing and insurance activities through improper billing and coding—some of which constitute medical fraud. While the rules are sufficiently complex that errors are inevitable, prior work has shown that some clinicians and healthcare institutions actively engage in ‘upcoding’ of patients’ diagnoses or severity to receive higher payments ^25–28^. Other healthcare entities, such as skilled nursing facilities, have also been observed to be engaged in this practice through “padding” of therapy times to increase revenues ^29^. The substantial national economic burden of billing and insurance activities and the specific contribution of variation in coding practices represents a pressing need for improvement. Recent work has attempted to take a surveillance approach through screening for outlier behavior in coding submissions at the institutional level ^30^. Still, we have not identified an implemented automated decision-support system that provides clinicians with coding guidance at the point of care. An NLP-based decision support tool for outpatient clinic notes might be feasible for mitigating clinician-driven administrative errors and costs associated with billing and insurance processes. However, careful attention is needed to ensure that such models are used to improve billing efficiency without further assisting upcoding

De-identification of clinical data is an active area of research given the risks of inadvertent release of PHI with data sharing in clinical and research contexts ^31^. A trade-off exists in supplying sufficient informative data versus revealing compromising PHI. The secondary objective of this study was to investigate the impact of clinic note de-identification on document classification performance. The de-identified note corpus showed a markedly lower classification accuracy compared to the model trained on the identified notes. This suggests that removing protected health information (PHI) elements from the clinical notes significantly affects the performance of the classifier model in assigning E/M LoS CPT codes. This was an unexpected finding as prior work has demonstrated that de-identifying clinical notes minimally reduces information ^32^. One possible explanation for this performance drop is a loss of contextual information and discriminative features. De-identification processes may inadvertently remove or obscure relevant contextual information that contains predictive features. It is possible that the identified data model was relying on “shortcut” features (i.e., learning to associate specific clinicians, specialty designation, or departments with specific coding patterns) ^33^. Such shortcut features could relate to the prediction target (i.e, the LoS E/M code) through one or more causal paths ^33^. Similarly, the de-identification process might introduce alterations to the natural flow and structure of the text that the model interprets. Another study trained on clinical notes from one emergency department setting found that the performance of word-embedding (WE)-based deep learning models did not differ when trained with identified and deidentified notes ^34^. It is plausible that the transformer-based models used in this study may rely more heavily on contextual information. Differing approaches in de-identification may also be a factor. Various methods and algorithms are available to de-identify clinical data ^35, 36^. Further research is needed to understand the potential implications of de-identification strategies on model performance across different NLP model architectures and de-identification approaches.

This study has several limitations which should be considered. First, the study was conducted using retrospective data from a single health system, which may limit the generalizability of our findings. Further research is needed to validate the performance of our NLP classifiers in different healthcare settings and patient populations, and it should be prospectively validated in other settings. Second, our classifiers were trained on specific specialties and subspecialties, and their applicability to other medical domains remains to be investigated. Third, the performance of the NLP classifiers is influenced by the quality and structure of the clinical notes, as observed by the variability in the model performance.

An additional validation step would be to assess the model’s performance against human billing auditors. Additionally, integrating the NLP-based decision support tool into electronic health record systems for seamless use by clinicians and billing staff might maximize its utility once validated. Last, estimating the economic and operational impact of the tool on healthcare costs, administrative workload, and potential reduction of billing errors and fraud may provide additional value to encourage implementation.

In conclusion, we found that NLP-based document classifiers could accurately predict E/M LoS CPT codes using clinical notes from various medical and procedural specialties. This could reduce the costs associated with this process. The models’ observed accuracy suggests that the classification task’s complexity merits further investigation. Our de-identification experiment demonstrated markedly lower classifier performance, suggesting that de-identification may negatively impact clinical documentation processing performance. This an important finding since de-identification is an emerging method for de-risked data sharing, and collaborative research is likely to be used more widely.

## METHODS

This study protocol was reviewed by our institutional review board and deemed exempt from formal review (Protocol #2021P002787). The development and reporting of this predictive model were completed following published guidelines from a multidisciplinary panel on the predictive model reporting ^37^.

### Setting and Prediction Goal

We developed natural language processing (NLP) document classifiers that assign evaluation and management (E/M) level of service (LoS) Current Procedural Terminology (CPT) billing codes to outpatient clinic notes. Our NLP classifiers were trained on retrospective clinical notes from a quaternary healthcare system. The success criteria for the multi-class classification tasks were the classification accuracy and weighted F1-score on internal test data. A secondary prediction goal was determining if clinic note de-identification impacted document classification performance.

### Dataset Development

To account for variations in coding practices among medicine, medicine-procedural, and surgical subspecialties, retrospective outpatient office clinic notes were selected from different medical specialties and subspecialties within Cardiology, Internal Medicine, Gastroenterology, and Otolaryngology-Head & Neck Surgery. Notes were obtained from clinic encounters spanning January 1, 2021 through December 31, 2021 to incorporate the latest reformed E/M LoS CPT coding definitions and criteria implemented by U.S. Centers for Medicare & Medicaid Services effective January 1, 2021 ^38^.

Clinic notes were included for ‘new’ patient encounters as defined by the usage of a new patient CPT code (i.e., CPT codes 99201–99205). The notes represent a wide range of patient (e.g., chief complaint, diagnosis, age) and clinician (e.g., providers, provider types (physician, physician-extender, nurse practitioners), hospital-based or community clinic sites) contexts. The LoS CPT code submitted previously for billing served as the ground truth label for each note. CPT codes corresponding to the five LoS strata were included (i.e., 99201– 99205 for new patients). Individual notes were excluded if a ground-truth LoS CPT code label was unavailable or if the note text was missing. Pre-processing the clinic notes dataset included removing infrequent labels.

### Clinical Note De-Identification

To remove protected health information (PHI) from each clinical note, we leveraged a pipeline that utilized a Named Entity Recognition (NER) model to annotate clinical notes for PHI elements (Spark NLP, John Snow Labs; Lewes, Delaware). Detected PHI elements included every instance of mentioned patient age, city, country, date, doctor/clinician name, hospital name, identification number, medical record number, health system organization/entity name, patient name, phone number, profession, state, street address number and name, username, and/or zip code. After identification, the PHI elements were masked in place using the type of element (**Table 1**).

### Model Development

We identified several state-of-the-art text classification models for these tasks. *Bio_ClinicalBERT,* a clinical language model pre-trained on EHR notes, was selected due to its demonstrated performance on clinical tasks ^39^. We also fine-tuned two general domain text classification models, *DistilBERT* ^40^ and *XLNet* ^21^. *DistilBERT* was chosen because it represents a smaller model that can run computationally constrained environments. *XLNet* was chosen as it has performance improvements over the *BERT* architecture ^21^. As clinic notes can be longer than 512 tokens, we also fine-tuned the *Clinical-Longformer* ^41^ model, which can handle an input sequence length of up to 4,096 tokens.

We fine-tuned each model on all the clinic notes and separately on each subspecialty note source. Model performance on the test set was determined by clinic note classification accuracy. We also computed weighted F1 scores to account for the imbalance of the classes. The F1 score combines the precision and recall of a classifier into a single metric. The weighted F1 score is the F1 score for each class weighted by its proportion in the dataset. Weighted F1 is useful in settings where an assessment of overall model performance is desired while accounting for the class imbalance. This analytic approach was repeated to serve the secondary objective using de-identified clinic notes for model fine-tuning.

For each fine-tuning experiment, the relevant clinic notes dataset was divided into 80% for model fine-tuning and 20% for testing. Model-specific tokenizers were used, and notes were padded to the longest sequence in the batch to ensure consistent input sequence lengths. Notes that exceeded the maximum token length of the model were truncated. Default fine-tuning training arguments were used across all experiments, including a learning rate of 2 x 10-5, a batch size of 3 due to memory constraints, five epochs, and a 0.01 weight decay.

### Computing Environment

Data processing and NLP modeling were completed in Python (vers. 3.9) and PyTorch (vers 1.13.1). We utilized the HuggingFace transformers hub to source the models and adapt the pre-trained model fine-tuning pipeline (available at: https://huggingface.co/). Models were trained in a Linux environment with one NVIDIA T4 GPU with 16GB of memory.

## Data Availability

As the raw data used in the present study contains PII/PHI, it is not available for public dissemination.

## Data availability

The data supporting this study’s findings are not publicly available due to the datasets containing protected health information (PHI) that could compromise research participant privacy.

## ACKNOWLEDGEMENTS

Dr. Crowson’s effort is partly supported by an NIH grant (Biomedical Informatics and Data Science Research Training Program; T15LM007092-30; PI Nils Gehlenborg). The authors thank and acknowledge John Snow Labs/Spark NLP for providing an academic research license for the de-identification toolkit.

