## Supplemental Tables for "Towards Medical Billing Automation: NLP for Outpatient Clinician Note Classification"

**S1.** Macro F1-scores for prediction of LoS E/M codes across general domain and clinical language models finetuned on notes from different medical specialties. All metrics are reported on test set data.

|  |  | **Macro F1-Score** | | | |
| --- | --- | --- | --- | --- | --- |
|  | **Full Dataset** | **Clinic Note Source** | | | |
| **Model** |  | *Cardiology* | *Gastroenterology* | *Internal Medicine* | *Otolaryngology-HNS* |
|  | *(n = 6,223)* | *(n = 2,656)* | *(n = 1,060)* | *(n = 2,036)* | *(n = 440)* |
| *Bio_ClinicalBERT* | 0.68 | **0.64** | **0.68** | 0.56 | 0.79 |
| *Clinical-Longformer* | 0.67 | 0.60 | 0.67 | 0.44 | **0.82** |
| *DistilBERT* | 0.65 | 0.62 | 0.65 | 0.55 | 0.81 |
| *XLNet* | **0.69** | 0.61 | 0.67 | **0.58** | 0.77 |
| *Mean (CI-95)* | 0.67 (0.65-0.69) | 0.62 (0.60-0.63) | 0.67 (0.65-0.68) | 0.53 (0.47-0.59) | 0.80 (0.78-0.82) |

**S2**. Class-specific F1 scores for each fine-tuned *Bio_ClinicalBERT* model

| **E/M Code** | **Full Dataset** | **Full Dataset / Deidentified** | **Cardiology** | **Gastroenterology** | **Internal Medicine** | **Otolaryngology** |
| --- | --- | --- | --- | --- | --- | --- |
| *Level II* | 0.48 | 0.0 | *N/A* | *N/A* | 0.37 | *N/A* |
| *Level III* | 0.70 | 0.41 | 0.30 | 0.44 | 0.71 | 0.88 |
| *Level IV* | 0.77 | 0.60 | 0.77 | 0.87 | 0.70 | 0.70 |
| *Level V* | 0.76 | 0.25 | 0.84 | 0.64 | 0.46 | *N/A* |

*N/A: no data*

**S3**. Class-specific F1 scores for each fine-tuned *Clinical-Longformer* model

| **E/M Code** | **Full Dataset** | **Full Dataset / Deidentified** | **Cardiology** | **Gastroenterology** | **Internal Medicine** | **Otolaryngology** |
| --- | --- | --- | --- | --- | --- | --- |
| *Level II* | 0.46 | 0.0 | *N/A* | *N/A* | 0.09 | *N/A* |
| *Level III* | 0.70 | 0.37 | 0.21 | 0.51 | 0.71 | 0.89 |
| *Level IV* | 0.77 | 0.61 | 0.77 | 0.87 | 0.68 | 0.74 |
| *Level V* | 0.78 | 0.13 | 0.82 | 0.63 | 0.26 | *N/A* |

*N/A: no data*

**S4**. Class-specific F1 scores for each fine-tuned *DistilBERT* model

| **E/M Code** | **Full Dataset** | **Full Dataset / Deidentified** | **Cardiology** | **Gastroenterology** | **Internal Medicine** | **Otolaryngology** |
| --- | --- | --- | --- | --- | --- | --- |
| *Level II* | 0.38 | 0.0 | *N/A* | *N/A* | 0.36 | *N/A* |
| *Level III* | 0.69 | 0.37 | 0.26 | 0.41 | 0.71 | 0.89 |
| *Level IV* | 0.77 | 0.61 | 0.77 | 0.88 | 0.69 | 0.73 |
| *Level V* | 0.75 | 0.16 | 0.83 | 0.66 | 0.42 | *N/A* |

*N/A: no data*

**S5**. Class-specific F1 scores for each fine-tuned *XLNet* model

| **E/M Code** | **Full Dataset** | **Full Dataset / Deidentified** | **Cardiology** | **Gastroenterology** | **Internal Medicine** | **Otolaryngology** |
| --- | --- | --- | --- | --- | --- | --- |
| *Level II* | 0.51 | 0.0 | *N/A* | *N/A* | 0.50 | *N/A* |
| *Level III* | 0.70 | 0.39 | 0.26 | 0.49 | 0.70 | 0.88 |
| *Level IV* | 0.77 | 0.59 | 0.75 | 0.87 | 0.70 | 0.66 |
| *Level V* | 0.76 | 0.25 | 0.81 | 0.64 | 0.43 | *N/A* |

*N/A: no data*
